# Quantifying the relationship between time to command-following and outcomes after TBI: the “1%” rule

**DOI:** 10.1101/2024.06.04.24308423

**Authors:** Samuel B. Snider, Hansen Deng, Flora M. Hammond, Robert G. Kowalski, William C. Walker, Ross D. Zafonte, David O. Okonkwo, Joseph T. Giacino, Ava M. Puccio, Yelena G. Bodien

**Author notes:** **Corresponding author** Samuel B. Snider, 60 Fenwood Road, Boston MA 02115. **Funding:** Dr. Snider receives funding from the National Institute of Neurologic Disorders and Stroke (1K23NS136767-01), American Heart Association, and National Institute of Biomedical Imaging and Bioengineering (1U01EB034228-01). Dr. Giacino receives funding from National Institutes of Health, National Institute of Neurologic Disorders and Stroke (U01 NS1365885; UG3NS117844-02; 5U01NS114140; 5UH3NS112826-04; 5R01NS102574-04; 5UH3NS095554-04), National Institute on Disability, Independent Living, and Rehabilitation Research (90DPCP0008; 90DPTB0011; 90DPHF0006; 90DPTB0027), and U.S. Department of Defense (W81XWH2210925; W81XWH-15-9-0001; W81XWH1910861). Dr. Hammond receives funding from National Institute on Disability, Independent Living, and Rehabilitation Research (grants 90DPTB0035, 90DPTB0022, 90DPTB0002, 90DPHF0006, 90DPTB0017, and 90RTEM0008); National Institutes of Health (UG3NS117844 and 1R01NS118009), PCORI UWSC9923/PCS-1604-35115; Department of Defense (W81XWH-18-1-0796); University of California-San Francisco; University of Michigan (SUBK10416CSPR-002). Dr. Kowalski receives funding from: NIH National Institute of Neurological Disorders and Stroke (LRP). Dr. Zafonte receives funding from National Institute on Disability, Independent Living and Rehabilitation Research (NIDILRR), Administration for Community Living (90DPCP0008-01-00, 90DP0039) Dr. Bodien receives funding from: NIH National Institute of Neurological Disorders and Stroke (U01 NS1365885, U01-NS086090), and the National Institute on Disability, Independent Living and Rehabilitation Research (NIDILRR), Administration for Community Living (90DPCP0008-01-00, 90DP0039). Dr. Walker receives funding from National Institute on Disability, Independent Living and Rehabilitation Research (NIDILRR), Administration for Community Living (90DBTB0021).

## Abstract

**Importance:** Recovery of command-following after traumatic brain injury (TBI) is an important prognostic indicator, however, the relationship between time to command-following and long-term functional outcome is not clear.

**Objective:** Evaluate the association between command-following and outcome 1-year after TBI. **Design** Cohort study of participants with moderate-severe TBI in the TBI Model Systems (TBIMS) who were followed 1-year post injury, and validation in an independent dataset from the Brain Trauma Research Center (BTRC) database.

**Setting:** TBIMS is a multi-center study of participants with moderate-severe TBI treated in an inpatient rehabilitation hospital. The BTRC database is derived from a single US level 1 trauma center and includes patients with severe TBI.

**Participants:** TBIMS: N=9,052 (mean±SD age 38±18 years, 76% male, 67% white); BTRC: N=228 (mean age 37±17 years, 76% male, 91% white). Participants did not follow commands on acute hospital admission and survived to discharge.

**Exposure:** Days to command-following during hospitalization.

**Main Outcome:** Glasgow Outcome Scale Extended (GOSE) score <4 (i.e., death or dependency) 1-year post TBI.

**Results:** Participants in TBIMS were more likely than those in BTRC to follow commands during acute hospitalization (90% vs 63%; p<0.001) and had a shorter median time to command-following (5 vs 9.5 days; p< 0.001). For each additional week without command-following, the odds ratio for death or dependency at 1 year was 1.30 (95% CI: [1.26,1.35]; p<0.001) in TBIMS and 1.49 ([1.15, 1.97]; p=0.003) in BTRC. Time to command-following had an AUC of 0.61 [0.59, 0.63] in TBIMS and 0.65 [0.53, 0.76]) in BTRC. Each additional day without command-following was associated with a 1.18% (1.16%, 1.20%) increase in the proportion of participants with death or dependency at 1-year in TBIMS and 1.05% (0.99%, 1.11%) in BTRC.

**Conclusion:** Time to command-following after moderate-severe TBI is associated with 1-year outcomes, but the predictive accuracy of absence of command-following on any single post-injury day is limited. In two independent cohorts, the likelihood of death or dependency increased by ∼1% for each additional day without command-following. Clinicians should be cautious when prognosticating based on the absence of command-following in the first five weeks after TBI.

## INTRODUCTION

After a traumatic brain injury (TBI), acute care clinicians often consider the persistent absence of behavioral responses to verbal instructions (i.e., command-following) to be an indicator of poor prognosis^1-3^. However, the accuracy of prognoses based on the absence of command-following at specific post-injury timepoints is unknown.

We used two large prospective TBI studies, the TBI Model Systems (TBIMS) National Database^4^ and the University of Pittsburgh Brain Trauma Research Center (BTRC) Database, together^5^ including more than 19,000 participants, to quantify the relationship between time to command-following and 1-year functional outcome.

## METHODS

### Study Cohorts

Institutional Review Boards at each TBIMS site and the BTRC approved the study, and participants’ surrogates provided informed consent. Characteristics of the TBIMS National Database^6, 7^ and the BTRC TBI study^5^ have been described previously. Briefly, TBIMS includes participants >16 years old with moderate-severe TBI who are admitted to TBIMS inpatient rehabilitation hospitals. The BTRC database includes participants aged 16-80 with severe TBI (admission Glasgow Coma Scale [GCS] score ≤ 8 with motor GCS ≤ 5) admitted to a level 1 trauma center. The BTRC database excludes patients with GCS total score=3 and fixed/dilated pupils or imminent brain death. From each study, we included participants who: 1) were >18 years old, 2) did not follow commands on the day of acute hospital admission, 3) survived acute hospitalization, and 4) were followed on the Glasgow Outcome Scale Extended [GOSE] 1-year post-TBI (TBIMS: N=9,052; BTRC: N=228; Table1, Figure 1).

**Figure 1:**
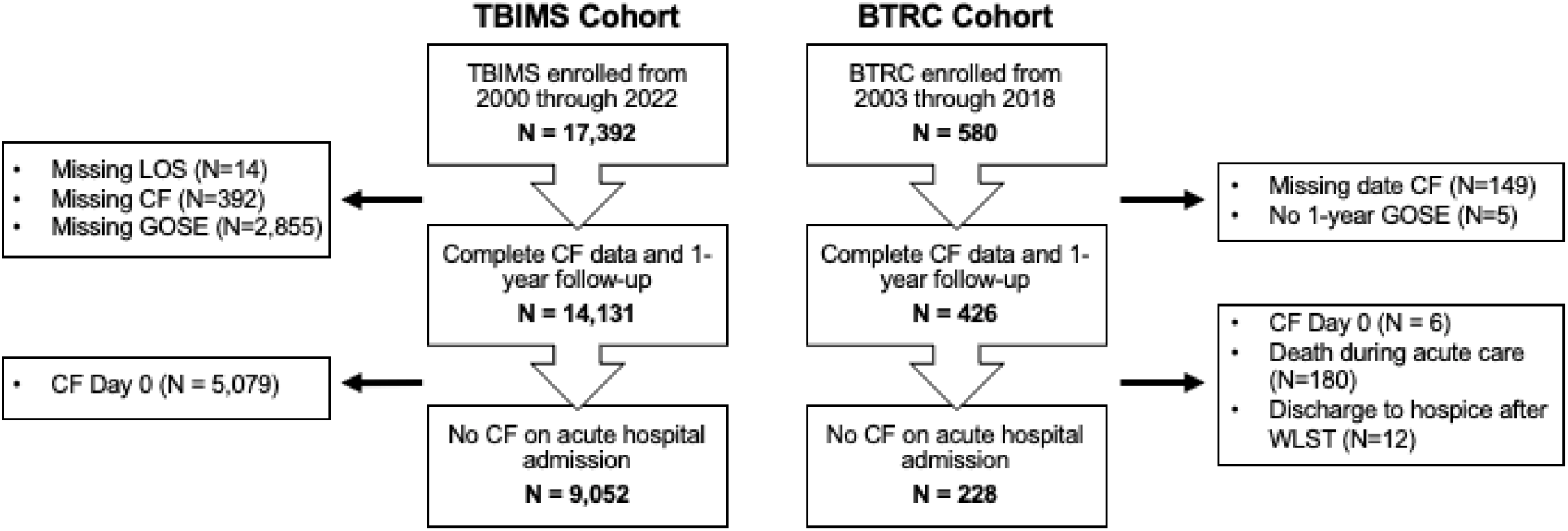
Study Flowchart. Study CONSORT diagram. Abbreviations: CF = command following; LOS = length of stay; GOSE = Glasgow Outcome Scale Extended; WLST = withdrawal of life sustaining treatment

### Primary Outcome

The primary outcome was GOSE<4 (death, vegetative state, lower severe disability, i.e., “death or dependency”) 1 year post-TBI.

### Command-Following: Definition and Analysis

Trained study staff followed standard operating procedures to review medical charts and identify the date of command-following: in TBIMS as the first occurrence of command-following documented in two clinical notes within 24 hours, and in BTRC as the first occurrence of command-following documented in a physician note on two consecutive hospital days. The observation period for command-following spanned from acute hospital admission to discharge.

We used logistic regression (predictors: cohort [TBIMS vs BTRC], days to command-following, and their interaction) and area under the receiver operating characteristic curve (AUC) to quantify the association between weeks to command-following and 1-year death or dependency in participants who followed commands during the observation period.

### Absence of Command-following: Definition and Analysis

We computed the proportion of participants with a 1-year outcome of death or dependency among all participants who did not follow commands on or before each of the first 50 days following acute hospital admission. Participants were included in this analysis until they followed commands or were discharged from the acute hospital. In each cohort, we used linear models to estimate the increase in the proportion of participants with 1-year death or dependency for each additional day without command-following.

## RESULTS

### Cohort Characteristics

The TBIMS cohort included N=9,052 participants (mean±SD age 38±18 years, 6,841 [76%] male, 6,040 [67%] white; Table 1). The BTRC cohort included N=228 participants (age 37±16 years, 174 [76%] male, 207 [91%] white; Table 1). TBI was less severe in the TBIMS compared to the BTRC cohort (median [IQR] GCS 10 [4, 13] vs 6 [5, 7]; p<0.001). All TBIMS participants, and 70% of the full BTRC cohort, received inpatient rehabilitation (p<0.001). Among BTRC participants who did not follow commands during acute hospitalization, 43% received inpatient rehabilitation. The proportion of participants with death or dependency at 1-year was lower in TBIMS compared to BTRC (23% vs 42%; p<0.001; Table 1; Supplementary Figure 1).

**Table 1:**
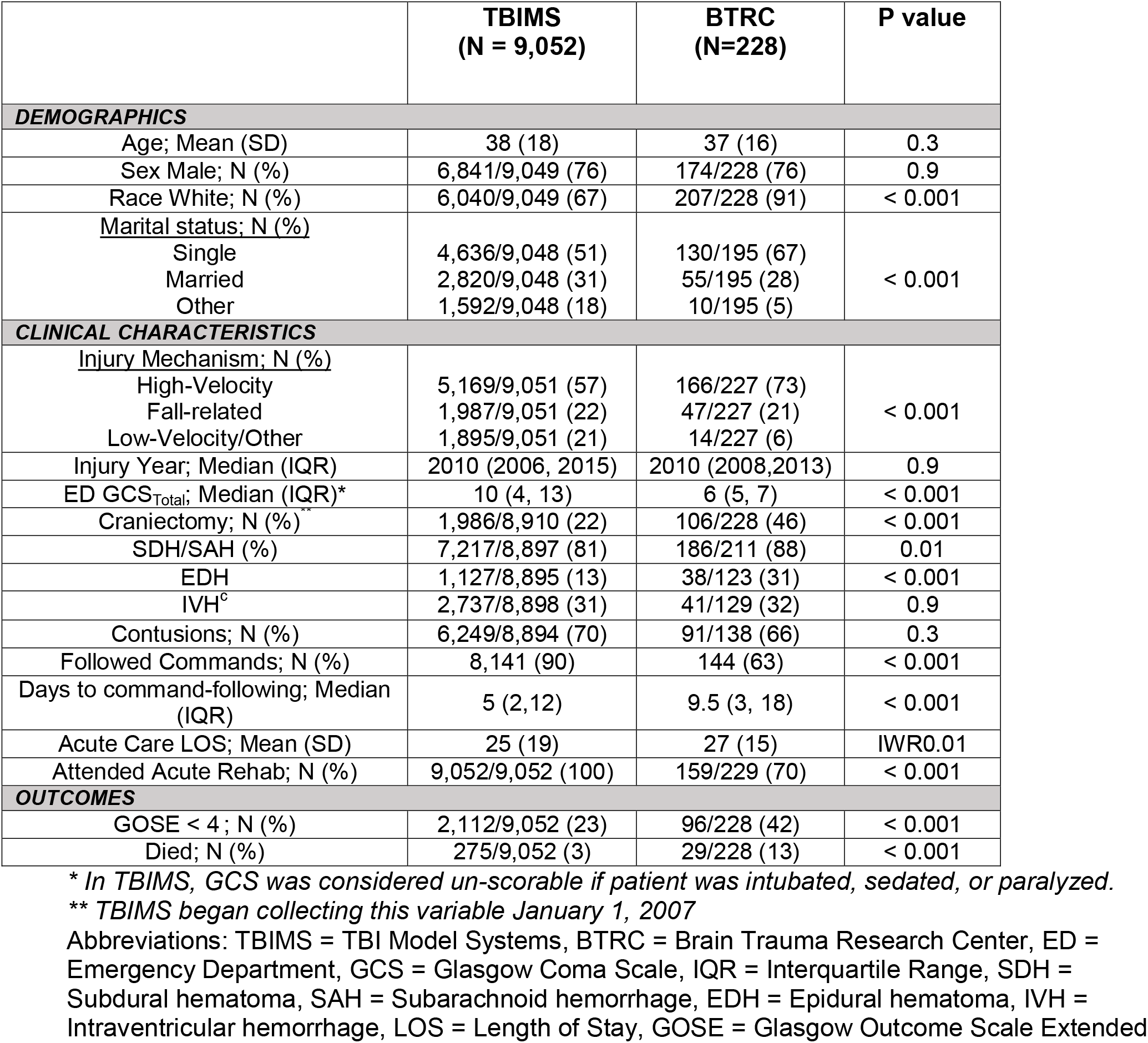
Cohort Characteristics

### Outcome in Participants who Followed Commands During Acute Hospitalization

During acute hospitalization, command-following was observed in 8,141 (90%) participants in TBIMS, and 144 (63%) in BTRC. Time to command-following was shorter in TBIMS compared to BTRC (median 5 days, IQR [2-12 days] vs 9.5 days [3-18 days]; p<0.001; Table 1, Figure 2A), but among command-followers, the proportion with 1-year death or dependency did not significantly differ between cohorts (20% [TBIMS] vs 22% [BTRC]; 𝒳^2^=0.3; p=0.6). Among participants who followed commands, the odds ratio (OR) for 1-year death or dependency with each additional week before command-following was 1.33 (95% CI: [1.29,1.38]; p <0.001) in TBIMS, with an AUC of 0.61 (95% CI: [0.59, 0.63]; Supplementary Figure 2) and 1.49 [1.15, 1.97]; p=0.003) in BTRC, with an AUC of 0.65 ([0.53, 0.76]; Supplementary Figure 2). The association between time to command-following and outcome did not differ between cohorts (cohort x command-following interaction *β*: -0.1 [-0.4, 0.2]; p=0.4).

**Figure 2:**
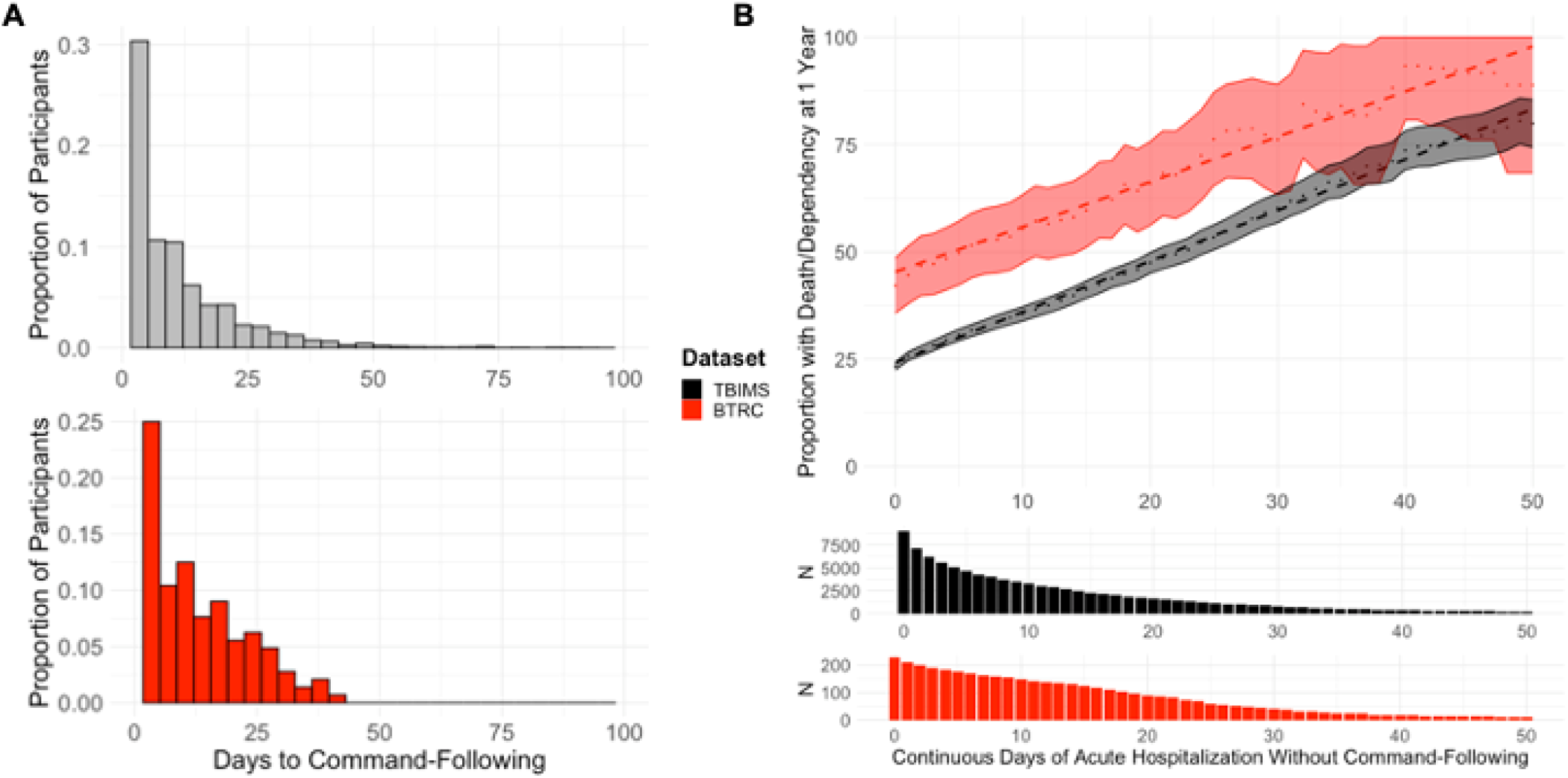
Command-following and 1-year Outcomes. (A): Proportion of participants whose first documented command-following occurred on each day starting from acute hospital admission (Day 0) through day 100 in TBIMS (black) and BTRC (red). (B Top): The proportion of participants with 1-year death or dependency (GOSE < 4, y-axis) is shown for those participants who did not follow commands on or before each day after acute hospital admission (TBIMS: black; BTRC: red). The dashed line represents the line of best fit of the daily proportions (individual data points). The shaded region represents the 95% confidence interval of the daily proportions calculated using the formula for the standard error of a proportion. (B lower panel): The histogram shows the total number of participants in each cohort who did not follow commands on or before each day following acute hospital admission.

### Outcome in Participants who Did Not Follow Commands During Acute Hospitalization

Command-following during acute hospitalization was not observed in 911 (10%) TBIMS and 84 (37%) BTRC participants. Compared to participants who followed commands at any point during acute hospitalization, those who did not had a higher incidence of death or dependency at 1 year (52% vs 20% [TBIMS]; OR 4.4 [3.8, 5.0], p<0.001; 76% vs 22% [BTRC]; OR 11.2 [6.0, 21.6], p<0.001). Each additional day without command-following was associated with a 1.18% (95% CI [1.16, 1.20]) increase in the proportion of participants with death or dependency in TBIMS (Figure 2B; black) and 1.05% [0.99, 1.11] increase in BTRC (Figure 2B; red).

## Discussion

In two longitudinal TBI cohorts, time to command-following had limited prognostic utility, discriminating weakly between GOSE<4 and GOSE≥4 1-year outcomes. Each additional hospital day without command-following yielded only a ∼1% increase in the proportion of participants with 1-year death or dependency– a rate that was remarkably consistent between cohorts. A high likelihood of death or dependency (i.e., > 90%) was observed only in participants who failed to follow commands for 40 or more days, which occurred rarely. These findings suggest that clinicians should avoid confidently assigning a poor prognosis based on the failure to recover command-following within the first 5 weeks post-TBI.

BTRC participants who never followed commands during acute care were more likely than TBIMS participants to have a 1-year outcome of death or dependency. This discrepancy is unsurprising given fundamental differences between the databases. While the BTRC includes a broad cohort of patients with severe TBI admitted to a level 1 trauma center, TBIMS includes only participants qualifying for and receiving inpatient rehabilitation. This may bias the TBIMS sample towards participants with a less severe injury, fewer medical comorbidities, and adequate insurance coverage^8^. Indeed, only 43% of BTRC participants who were not following commands at acute hospital discharge received inpatient rehabilitation.

A limitation of this study is that the date of command-following was determined by systematic chart review; monitoring frequency may have varied across subjects, and potential confounds (e.g., continuous sedation) were not recorded. In addition, to avoid the “self-fulfilling prophecy” bias^9^, we excluded participants who died during acute hospitalization due to their injuries or from withdrawal of life sustaining treatment. However, in doing so, we may have systematically excluded more-severely injured patients with a higher likelihood of death or dependency. Finally, due to sample size constraints, we were unable to evaluate outcomes in participants who were not following commands for more than 50 days after injury.

## Conclusion

Although time to command-following was associated with outcomes, in the first five weeks following TBI, each additional day without command-following was associated with only a 1% increase in the proportion of participants with death or dependency at 1 year. Our results support a cautious approach towards using the absence of command-following as a negative prognostic marker in the first five weeks after TBI.

## Data Availability

All data produced in the present study are available upon reasonable request to the authors

## SUPPLEMENTARY MATERIAL

**Supplementary Figure 1:**
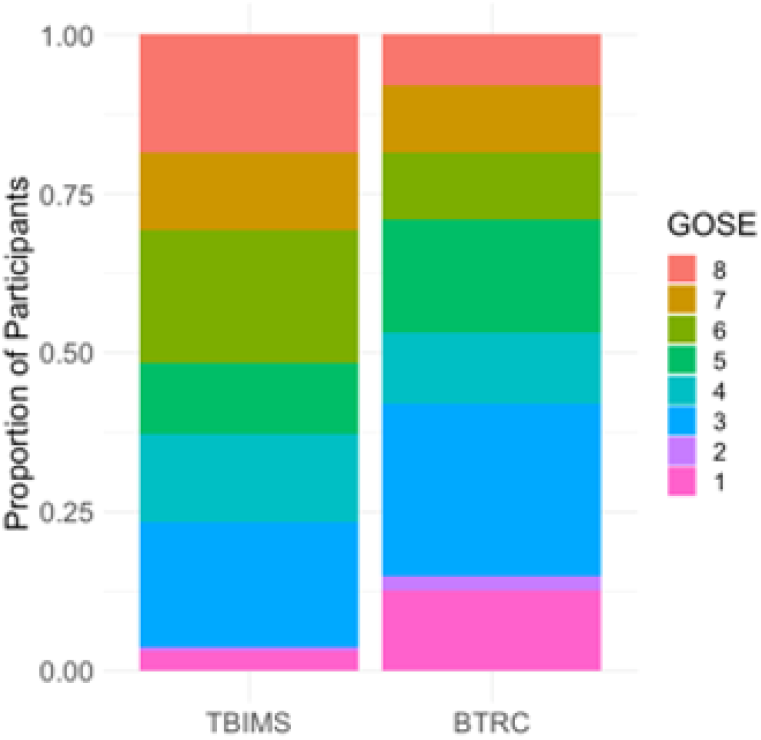
Proportion of GOSE Scores in Each Dataset. Proportion of participants in each GOSE category at 1-year following injury in TBIMS and BTRC. GOSE Categories: 1 = Death, 2 = Vegetative State, 3 = Lower Severe Disability, 4 = Upper Severe Disability, 5 = Lower Moderate Disability, 6 = Upper Moderate Disability, 7 = Lower Good Recovery, 8 = Upper Good Recovery

**Supplementary Figure 2:**
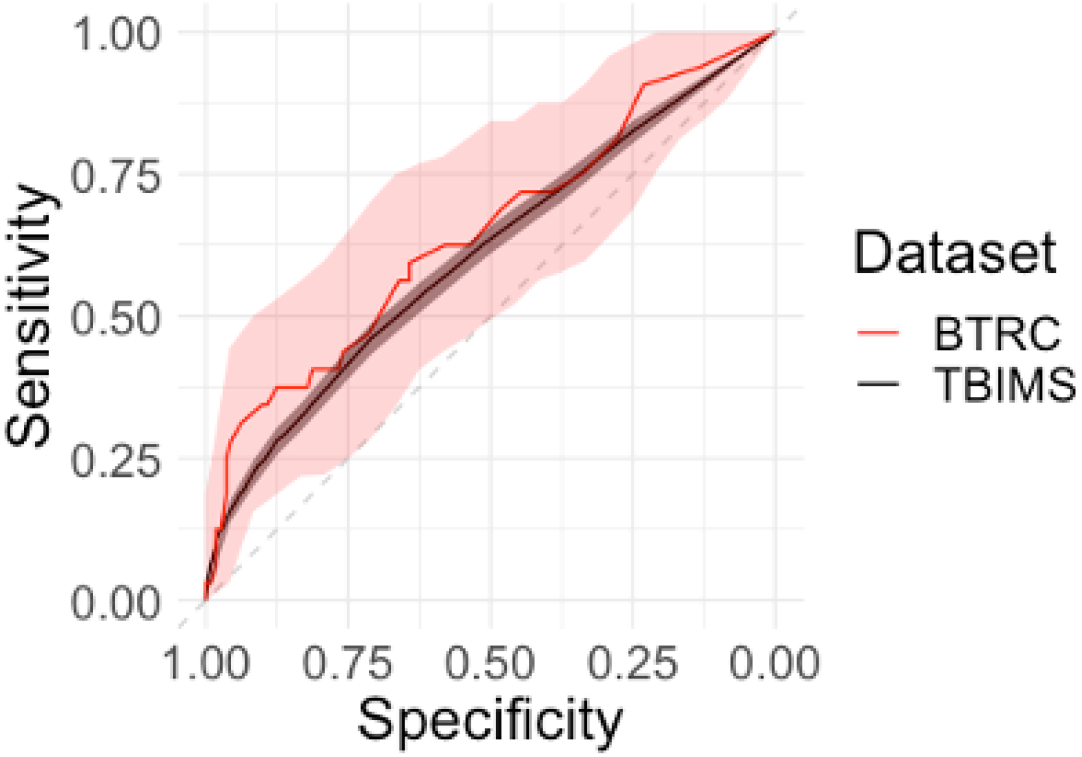
Time to Command-Following ROC Curves. ROC curves for time to command-following among participants who followed commands before acute care discharge in TBIMS (black) and BTRC (red).

## Notes

**Disclosures/Conflicts of Interest:** Dr. Hammond receives royalties from Springer/Demos and Lash Publishing, and she has served on Advisory Boards for Avanir and Otsuka Pharmaceuticals. Dr. Zafonte reported receiving royalties from Springer/Demos for the text Brain Injury Medicine as well as serving on the scientific advisory boards of Myomo, and One Care.ai. Dr. Giacino occasionally receives honoraria from academic and medical institutions for conducting training seminars on the Coma Recovery Scale-Revised.

### Competing Interest Statement

Dr. Hammond receives royalties from Springer/Demos and Lash Publishing, and she has served on Advisory Boards for Avanir and Otsuka Pharmaceuticals.
Dr. Zafonte reported receiving royalties from Springer/Demos for the text Brain Injury Medicine as well as serving on the scientific advisory boards of Myomo, and One Care.ai.
Dr. Giacino occasionally receives honoraria from academic and medical institutions for conducting training seminars on the Coma Recovery Scale- Revised.

### Author Declarations

Institutional Review Boards at each TBIMS site and the BTRC approved the study, and participants surrogates provided informed consent.

